# Latent Class Analysis of COVID-19 Deaths by Comorbidities— United States, February-May 2020

**DOI:** 10.1101/2024.04.16.24305728

**Authors:** David A. Siegel, William A. Bentley, Rongxia Li, Carolyn V. Gould, David W. McCormick, Joseph McLaughlin, Anjali Vyas, Charles R. Clark, Gillian Richardson, Anna Krueger, Stacy Holzbauer, Leslie Kollmann, Deepam Thomas, Mojisola Ojo, Marc Paladini, Kathleen H. Reilly, Meagan McLafferty, Dean E. Sidelinger, Laura C. Chambers, Jennifer S. Read, Mona Saraiya, Paul R Young, Jonathan M. Wortham, Emilia H. Koumans

**Affiliations:** CDC COVID-19 Emergency Response Team, Centers for Disease Control and Prevention, 4770 Buford Hwy NE, MS-76, Atlanta, GA 30341, USA; Alaska Department of Health and Social Services, 3601 C St #902, Anchorage, AK 99503, USA; Hawaii State Department of Health, 1250 Punchbowl St, Honolulu, HI 96813, USA; Indiana Department of Health, 2 N Meridian St, Indianapolis, IN 46204, USA; Louisiana Department of Health, 628 N 4th St, Baton Rouge, LA 70802, USA; Maine Center for Disease Control and Prevention, State House Station 11, 286 Water St, Augusta, ME 04333, USA; Minnesota Department of Health, P.O. Box 64975, St. Paul, MN 55164-0975, USA; New Jersey Department of Health, P.O. Box 360, Trenton, NJ, USA; New York City Department of Health and Mental Hygiene, 42-09 28th St; Long Island City, NY 11101, USA; Oregon Health Authority, 800 NE Oregon St, Portland, OR 97232, USA; Rhode Island Department of Health, 3 Capitol Hill, Providence, RI 02908, USA; Infectious Disease Epidemiology, Vermont Department of Health, 108 Cherry Street, Burlington, VT 05402, USA; Larner College of Medicine, University of Vermont, Given Medical Bldg, E-126, 89 Beaumont Ave, Burlington, VT 05405, USA

**Keywords:** COVID-19, Mortality, Latent Class Analysis

## Abstract

**Purpose:** Risk factors for coronavirus disease 2019 (COVID-19) mortality include older age, cardiovascular disease, diabetes, and other comorbidities. Latent class analysis (LCA) can identify unrecognized morbidity patterns for decedents with COVID-19.

**Methods:** Data were collected from 23 U.S. jurisdictions about decedents with COVID-19 early in the COVID-19 pandemic (February 12–May 12, 2020). LCA identified groups of individuals based upon pre-existing comorbidities: cardiovascular, renal, lung, neurologic, and liver disease; obesity; diabetes; and immunocompromised state. Results were stratified by sex, age, race/ethnicity, and location of death.

**Results:** Of 12,340 decedents, LCA identified three classes, which included classes with prominent cardiovascular disease and diabetes (32%), prominent cardiovascular disease without diabetes (19%), and a “minimal prevalence” class (49%) with a low frequency of comorbidities. Individuals in the “minimal prevalence” class had risk factors in <2 comorbidity groups, where cardiovascular disease was the most common for individuals with a single comorbidity.

**Conclusions:** LCA analysis reaffirms the importance of diabetes and cardiovascular disease as risk factors for COVID-19 mortality and indicates that about half of decedents were in the “minimal prevalence” group. Results could guide vaccination and treatment messaging to groups with no or few underlying conditions.

**Disclaimer:** The conclusions, findings, and opinions expressed by the authors do not necessarily reflect the offical position of the Centers for Disease Control and Prevention or the authors’ affililated institutions.

## Introduction

Numerous risk factors have been independently associated with increased risk of coronavirus disease 2019 (COVID-19)-related mortality; regression analyses have identified male sex,^1, 2^ older age,^2-9^ obesity,^6, 10^ cardiovascular disease,^4, 11^ diabetes,^8, 12^ and other comorbidities as being associated with death.^4, 13^ Despite these previous studies of risk factors for COVID-19-related mortality, unrecognized subgroups of individuals and their demographic and clinical characteristics may exist that could inform public health interventions, such as vaccination and direction of treatment, to prevent future deaths.

Latent class analysis (LCA) is a well-validated modeling technique that classifies individuals into subgroups based upon unobservable, unbiased patterns of disease manifestation and has not been widely utilized to describe patient characteristics with COVID-19.^14, 15^ The objective of this study is to use LCA to assess previously unobserved differences among decedents with COVID-19 and whether those groupings are associated with certain demographic characteristics.

## Materials and methods

The Centers for Disease Control and Prevention (CDC) solicited information on individuals who died with COVID-19 from state, territorial, and local public health departments.^16^ Among 56 public health departments contacted by CDC, 23 provided data as part of this request. Individuals were included if they died during February 12–May 12, 2020 and had been diagnosed with PCR-positive severe acute respiratory syndrome coronavirus 2 (SARS-CoV-2) infection.

Case data were abstracted from medical charts and death certificates by participating public health departments. Available demographic data included sex, age, race/ethnicity, and location of death (e.g., home, hospital). Comorbidities were categorized based upon recognized risk factors for mortality^4, 8, 16-18^ and included cardiovascular disease, diabetes, renal disease, chronic lung disease, immunocompromised state, neurologic disease, obesity, and liver disease. Specifically, comorbidities included cardiovascular disease (congenital heart disease, coronary artery disease, congestive heart failure, hypertension, cerebrovascular accident/stroke, valvular heart disease, conduction disorders or dysrhythmias, pulmonary hypertension, dyslipidemia, thrombosis, other cardiovascular disease), diabetes mellitus types I and II, chronic kidney disease (chronic kidney disease, end-stage renal disease, other kidney diseases), chronic lung disease (chronic obstructive pulmonary disease/emphysema, asthma, tuberculosis, other chronic lung diseases), immunocompromised state (cancer, human immunodeficiency virus [HIV] infection, lupus, ulcerative colitis, Crohn’s disease, identified as being immunosuppressed), neurologic conditions (dementia, seizure disorder, other neurologic conditions), obesity (body mass index ≥30 kg/m^2^), and chronic liver disease (cirrhosis, alcoholic hepatitis, chronic liver disease, end-stage liver disease, hepatitis B, hepatitis C, nonalcoholic steatohepatitis, other chronic liver diseases). A decedent with comorbidities in the same group (e.g., heart failure and stroke) was only counted in that comorbidity group (e.g., cardiovascular disease) once. Presence of a comorbidity was determined through medical chart review to be yes, no, or unknown (including missing). Other comorbidities outside of these eight groups were not uniformly reported by participating jurisdictions and were therefore not included in this analysis.

LCA modeling was used to identify characteristics that clustered among decedents using comorbidity groups.^19^ To select the appropriate number of classes, two-, three-, and four-class models were first fit to the data. A three-class model was selected based on the Bayesian information criterion. Thirty replications of the model were run with a maximum of 10,000 iterations each to maximize the chance of finding a good global fit and avoid local maximums. Individuals were assigned exclusively to one of the three classes according to their highest probability of membership.^19^ P-values were calculated from Pearson’s chi-squared test to compare the distribution of variables. LCA was run for the entire study population and then separately for each age group. Classes were named after the 1 or 2 comorbidities with the highest prevalence or as “minimal prevalence” class where the prevalence of each comorbidity group was less than that of the entire sample. Data analysis was performed using R (version 4.0.3) and SAS (version 9.4, SAS Institute, Cary, NC); LCA was performed by the R package version 1.4.1.^20, 21^ This activity was reviewed by CDC and was conducted consistent with applicable federal law and CDC policy.^1^

## Results

Among 12,340 decedents with COVID-19, 60% were male, 75% were age 65 years or older, 32% were non-Hispanic White, 24% were non-Hispanic Black, and 21% were Hispanic (Table 1). Of decedents, 60% were from New York City, 18% were from New Jersey, 8% were from Washington state, and 14% were from other public health departments. Two-thirds (66%) of decedents died in the hospital, 7% in long-term care facilities, 5% in emergency departments, and for 12% location of death was unknown. The most common comorbidities were cardiovascular disease (58%), followed by diabetes (36%), renal disease (20%), and chronic lung disease (18%).

**Table 1:**
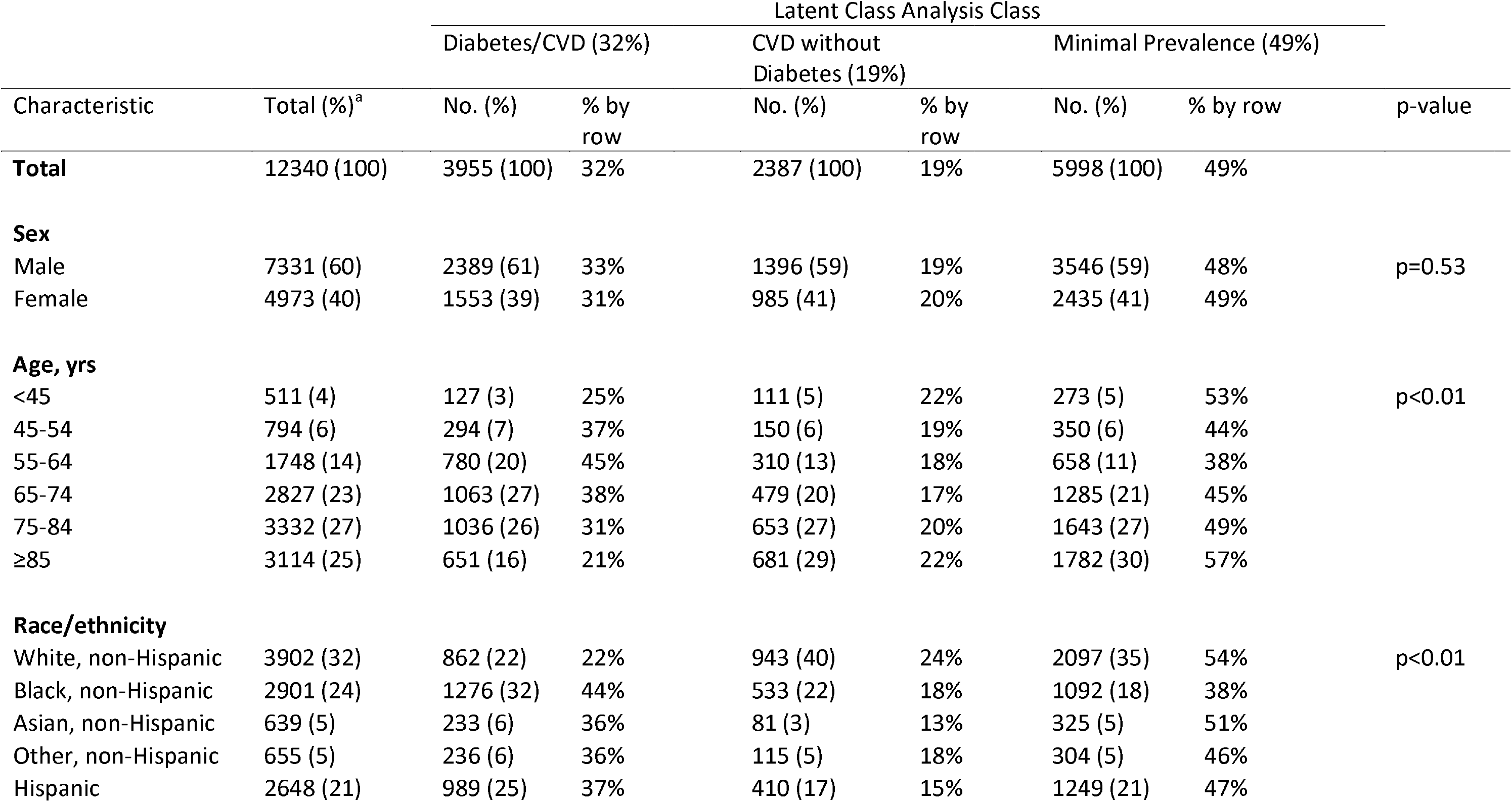
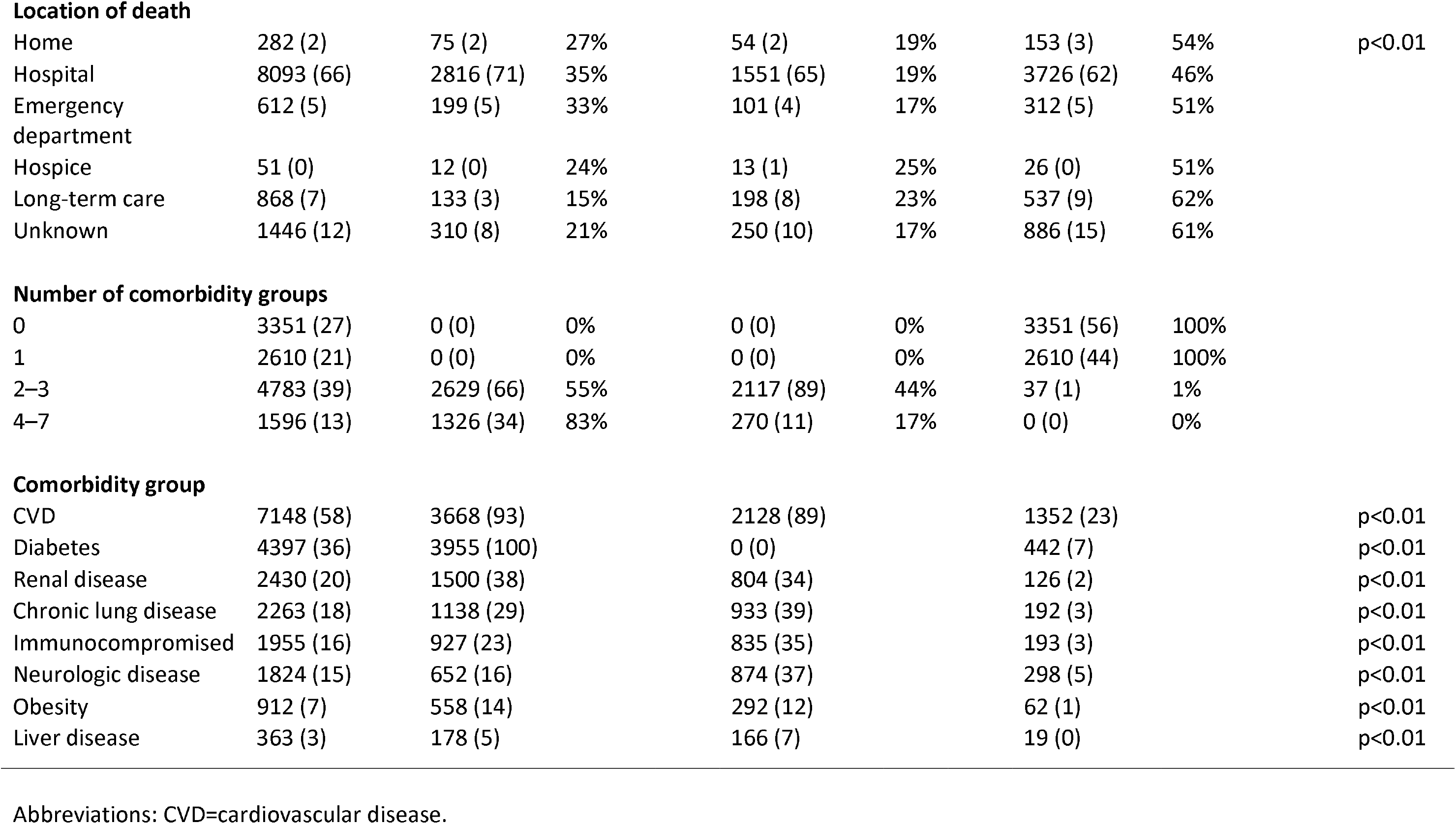
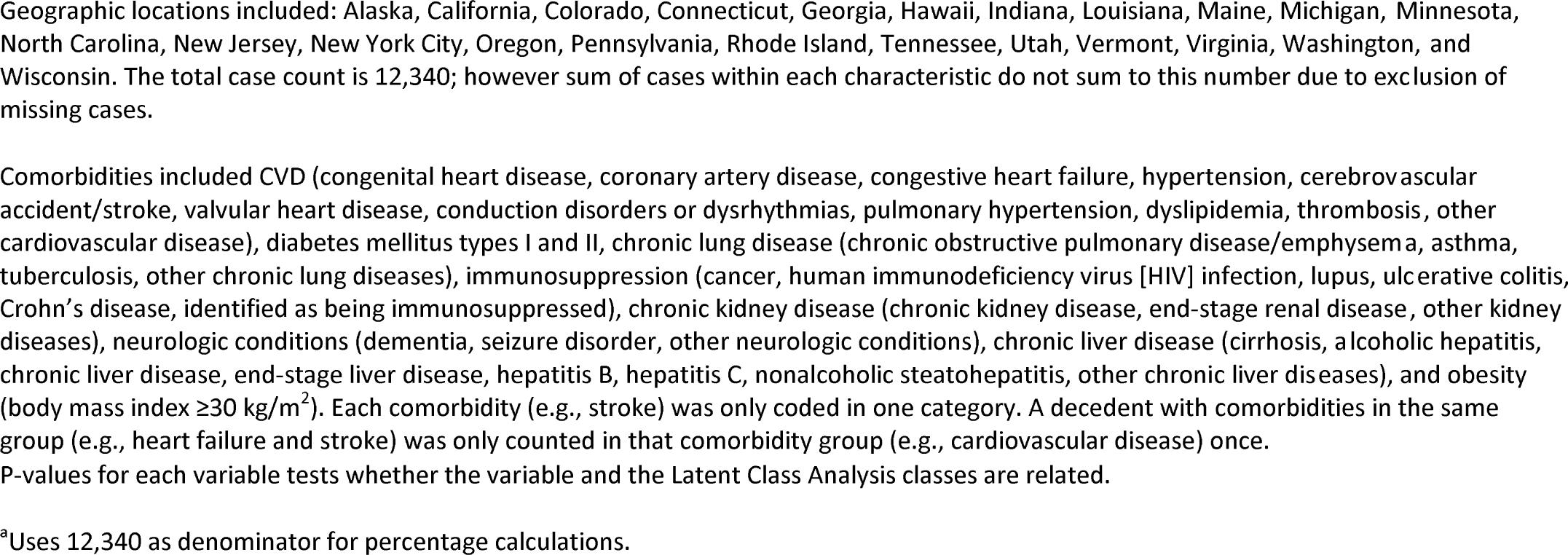
Demographic characteristics and latent class analysis of deaths with COVID-19, February 12–May 12, 2020.

LCA of all ages combined identified three classes of individuals (Table 1): a “diabetes and cardiovascular” class (n=3955, 32% of decedents), a “cardiovascular without diabetes” class (n=2387, 19%), and a “minimal prevalence” class (n=5998, 49%). In the “diabetes and cardiovascular” class, 100% of decedents had diabetes, 93% had cardiovascular disease, and 38% had renal disease. In the “cardiovascular without diabetes” class, 89% of decedents had cardiovascular disease, 39% had chronic lung disease, and 37% had neurologic disease. In the “minimal prevalence” class, which represented individuals with 0 or 1 comorbidity groups, 23% of decedents had cardiovascular disease, and all other conditions had a prevalence of ≤7%.

Across age groups, the ≥85 years age group had the highest proportion of decedents grouped into the “minimal prevalence” class (57%). The “diabetes and cardiovascular” class was more common among non-Hispanic Black (44%) than non-Hispanic White (22%) decedents, and the “minimal prevalence” class was most common among non-Hispanic White (54%) and non-Hispanic Asian decedents (51%), compared with non-Hispanic Black decedents (38%).

When LCA was run separately for each age group, each age group consisted of a class with cardiovascular disease being the most prevalent comorbidity group, a class where the most prevalent comorbidity varied, and a “minimal prevalence” class where the prevalence of each comorbidity group was less than that of the entire sample (Table 2).

**Table 2:**
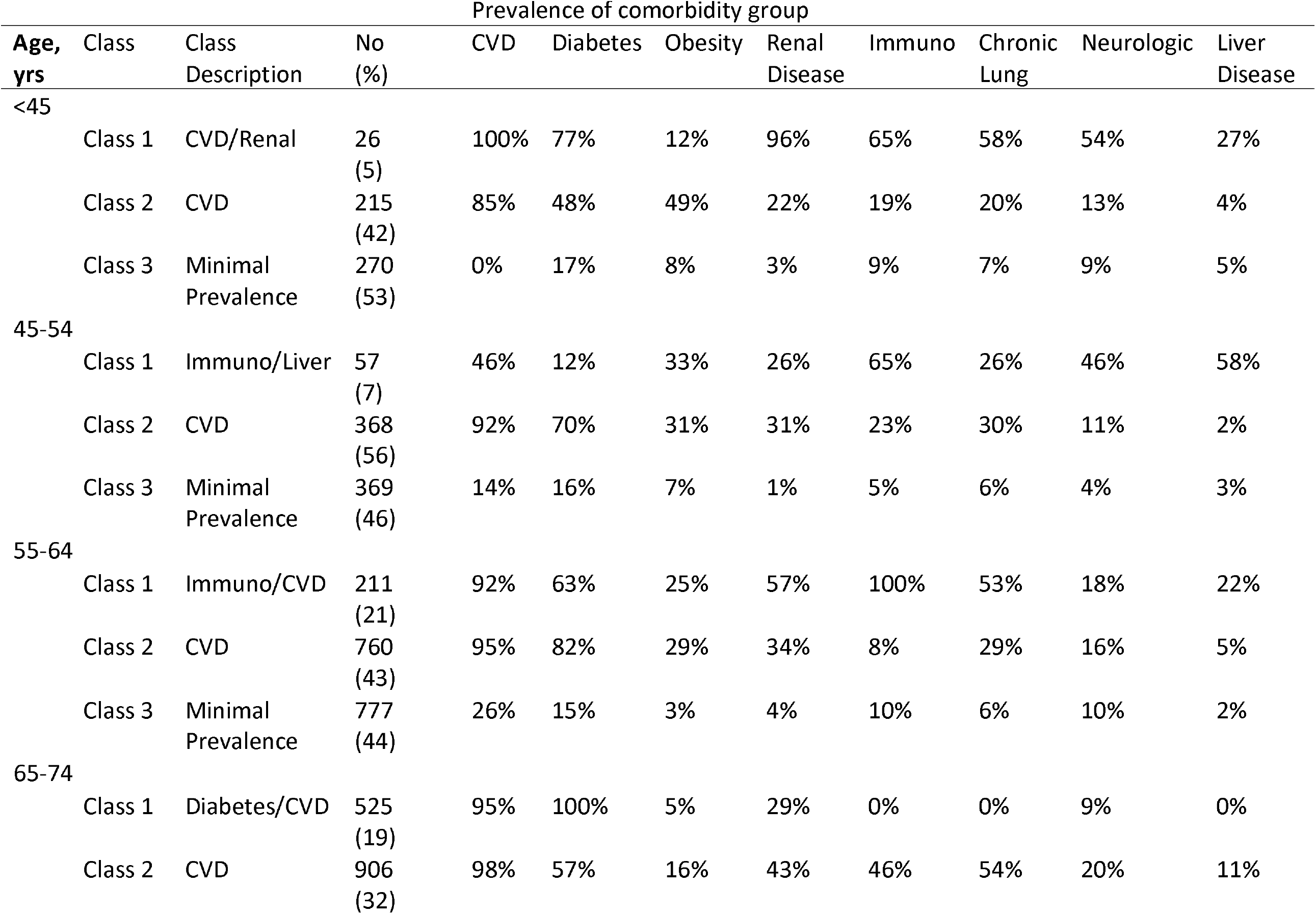
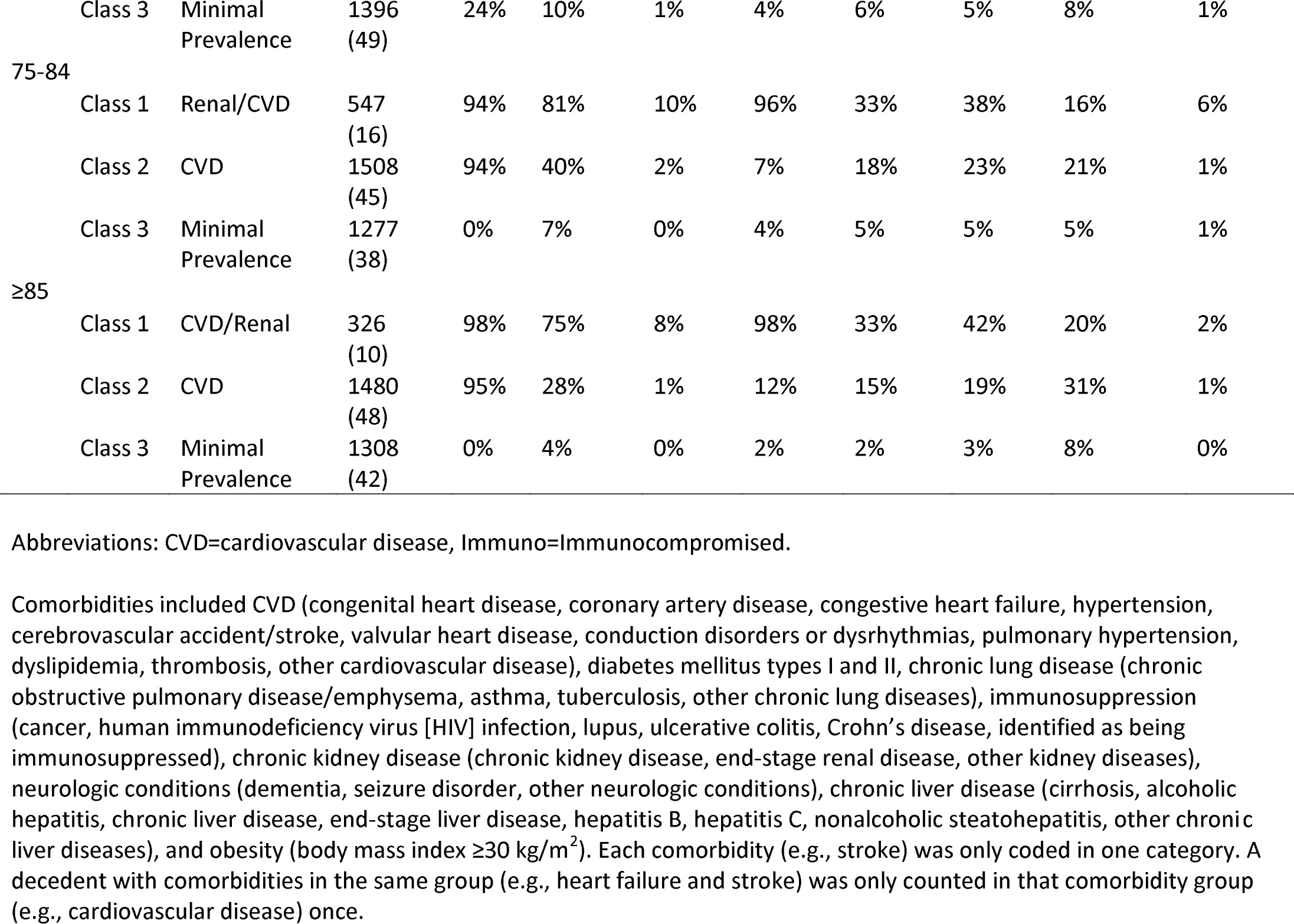
Latent class analysis by age and prevalence of comorbidity group among of deaths with COVID-19, February 12– May 12, 2020.

## Discussion

This study describes mortality patterns among individuals, mostly in the Northeast United States, early in the COVID-19 pandemic, and reflects patterns of death for individuals before antivirals or vaccination against SARS-CoV-2 was available. Nearly half of all decedents with COVID-19 could be subtyped into a class with a low frequency of measured comorbidities; this class represented individuals with 0 or 1 comorbidities, of which cardiovascular disease was the most common. Conversely, about half of decedents could be subtyped into a class with a predominant prevalence of cardiovascular disease or diabetes. Racial/ethnic differences were seen in the percentage of decedents in the “minimal prevalence” class, with non-Hispanic Whites having a higher proportion within this class among all non-Hispanic White decedents than the proportion of non-Hispanic Blacks who were within this class among all non-Hispanic Black decedents.

The presence of classes with a high prevalence of cardiovascular disease and diabetes indicates the importance of this morbidity pattern for individuals with COVID-19, consistent with prior literature.^4, 22-24^ Cardiovascular disease was present at 23% even in the “minimal prevalence” class, which might indicate the importance of this factor independent of other comorbidities. Other comorbidities such as renal, liver, and neurologic disease have been associated with higher COVID-19 mortality rates.^8^ Renal disease, which had a 20% prevalence in this study population, was the third most prevalent comorbidity in the “diabetes/cardiovascular” class. Chronic lung disease was the second most prevalent condition in the “cardiovascular without diabetes” class. Past studies have found chronic pulmonary or respiratory disease to be a risk factor for mortality.^11, 24^

Non-Hispanic Black and non-Hispanic White decedents had the lowest and highest percentages in the “minimal prevalence” class, respectively. Some studies have found that race is independently associated with a higher risk of death with COVID-19.^8, 9, 11, 25, 26^ Disparities among races and ethnicities in mortality with COVID-19 could be related to a number of factors such as prevalence of infection, presence of comorbidities, or societal factors.^27^ However, when adjusting for clinical and sociodemographic factors, race might not be an independent predictor of mortality with COVID-19.^25^

Older age is a well-established risk factor for mortality with COVID-19,^4^ which is reflected in the decedent age distribution of this study; 75% were age 65 years or older. Despite this age distribution, the presence of a “minimal prevalence” class with low frequencies of measured comorbidities persisted across all age groups, ranging from 57% of cases with ages ≥85 years to 38% of cases in the 55-64 years age group. The baseline prevalence of comorbidities by age in this report is consistent with other reports of age-specific baseline prevalence of comorbidities.^28, 29^ A large proportion (57%) of decedents aged 85 years or older were in the “minimal prevalence” class, which might be because age was their primary risk factor for death with COVID-19. Older decedents in this study might have had multiple comorbidities within the same group (e.g., stroke and heart failure), had more severe comorbidities, or had comorbidities not measured by this study (e.g., gastrointestinal, musculoskeletal, or psychiatric conditions).

This analysis has several limitations. First, this analysis represents a convenience sample of individuals who died with COVID-19 and represents approximately 15% of the 80,000 individuals who had died with COVID-19 nationwide by May 12, 2020.^30^ Additionally, because most of decedents in this study were from New York City and New Jersey, these findings mostly represent a Northeast, urban population and might not be generalizable nationally. Second, there might have been variation in SARS-CoV-2 testing protocols among health departments and resulting variation in case ascertainment. Third, there might have been variation in diagnosis and recording of comorbidities; some comorbidities might be present but undiagnosed, and recording of comorbidities might vary by source of case ascertainment (e.g., death certificate). Fourth, only eight comorbidity groups were represented in these data. Other pre-existing comorbidities might have been present and were unmeasured, for example, disabilities, which have been linked to higher risk of severe disease with COVID-19.^31^ Fifth, socioeconomic status was not collected, limiting the ability of assessing access to care and its association with mortality. Sixth, the study did not quantify the number of people included in the study who had COVID-19 but died from an unrelated cause. Finally, these data represent disease early in the pandemic and reflect patterns of death before interventions such as dexamethasone were widespread, early in the course of nationwide prevention efforts (e.g., mask mandates), emergence of variants, and COVID-19 vaccination.

Individuals with older age, cardiovascular disease alone, or cardiovascular disease in combination with diabetes, represent distinct subpopulations that could be targets for COVID-19 prevention efforts, such as mitigation measures and vaccination promotion. Conversely, decedents in the “minimal prevalence” class across all age categories largely did not have diabetes and cardiovascular disease; this group represented a subpopulation with mortality risk factors mostly independent from the eight comorbidities included in this report. However, cardiovascular disease was the most common single comorbidity. Communication directed to individuals in a potential “minimal prevalence” class could emphasize the importance of vaccination or antiviral therapy even for those without comorbidities such as diabetes and cardiovascular disease. Over two-thirds of individuals aged 18-25 years who were not planning on getting the COVID-19 vaccine cited it was because “I am not a member of a high-risk group.” Further evaluation of a potential “minimal prevalence” class could benefit messaging for individuals in this group.^32^ Alternatively, individuals in a potential “minimal prevalence” class could have undiagnosed risk factors for death with COVID-19; for these individuals, maintaining routine medical care could be helpful in identifying potential underlying conditions. Future research using latent class analysis could identify how additional factors may have contributed to severity and mortality clusters among individuals with COVID-19.

## Data Availability

Underlying data are not publicly available with personal identifiable information due to privacy and legal restrictions.

## Abbreviations

(CDC): Centers for Disease Control and Prevention
(COVID-19): Coronavirus disease 2019
(HIV): Human immunodeficiency virus
(LCA): Latent class analysis
(PCR): Polymerase chain reaction
(SARS-CoV-2): Severe acute respiratory syndrome coronavirus 2

## Acknowledgements

Sandy Althomsons, James Lee, Michigan Department of Health and Human Services, Lansing, MI, North Carolina Department of Health and Human Services, Raleigh, NC, Pennsylvania Department of Health, Harrisburg, PA, Wisconsin Department of Health Services, Madison, WI. We would additionally like to acknowledge staff from: California Department of Public Health, Colorado Department of Public Health and Environment, Connecticut Department of Public Health, Georgia Department of Public Health, Tennessee Department of Health, Utah Department of Health, Virginia Department of Health, and Washington State Department of Health.

## Footnotes

1 See e.g., 45 C.F.R. part 46.102(l)(2), 21 C.F.R. part 56; 42 U.S.C. §241(d); 5 U.S.C. §552a; 44 U.S.C. §3501 et seq.

